# A 12-month randomised, double-blind, controlled, multicentre trial comparing changes in Cigarette consumption after switchinG to high or low nicotine strENght E-cigaretteS In smokers with Schizophrenia spectrum disorders: Protocol for the GENESIS Trial

**DOI:** 10.1101/2020.10.15.20141457

**Authors:** Pasquale Caponnetto, Bulat Idrisov, Maria Salvina Signorelli, Evgeny Krupitsky, Tetiana Kiriazova, Ramin Nilforooshan, Fabio Cibella, Marilena Maglia, Daniela Saitta, Francesca Benfatto, Eugenio Aguglia, Roberto Cavallaro, Lucio Inguscio, Giuseppe Minutolo, Roberta Auditore, Riccardo Polosa, GENESIS study investigators

## Abstract

**Background:** Smoking prevalence among people with mental disorders is about two to four times higher than in the general population. As a result of high smoking rates, people with a mental health condition also have high rates of morbidity and mortality from smoking-related diseases compared with the general population. Progress in reducing smoking prevalence in people with mental health diagnoses has been very slow compared to the general population. Consequently, there is a pressing need for alternative and more efficient interventions to reduce or prevent morbidity and mortality in smokers with schizophrenia spectrum disorders.

**Methods:** A volunteer population of 258 adult smokers with Schizophrenia Spectrum Disorder will be recruited for the GENESIS study, a randomized, double blind, smoking cessation trial comparing effectiveness, safety and subjective effects between 5% and 1.5% nicotine e-cigarette. The study duration will be 12-month. The primary endpoint of this study will be the continuous quit rate defined as the proportion of study participants who self-report that they had stopped smoking at 6-month, biochemically verified by exhaled CO measurements of ≤ 7 ppm. These participants will be referred to as “Quitters”. The differences in continuous variables between the two groups for normally distributed data will be evaluated by one-way analysis of variance (ANOVA). The differences between the two groups for not normally distributed data will be evaluated by the Mann-Whitney U test. Any correlation between the variables under evaluation will be assessed by Spearman r correlation. To analyze differences in frequency distribution of categorical variables we will use the Chi-square test with the Yates correction or the Fisher exact test. All statistical tests are two-tailed and are considered to be statistically significant at a P value <0.05. The consistency of effects for pre-specified subgroups will be assessed using tests for heterogeneity. Subgroups will be based on age, sex, education, level of nicotine dependence.

**Discussion:** This will be the first multicenter randomized trial directly comparing high (JUUL 5% nicotine) with low nicotine strength devices (JUUL 1.5% nicotine) in term of reduction in cigarette consumption, adoption rates, product acceptability, tolerability, and tobacco harm reduction potential. This knowledge can contribute to a better understanding of e-cigarette with high nicotine content as a pragmatic and much less harmful alternative to tobacco smoking with the possibility of significant health gains in smokers with schizophrenia spectrum disorders.

**Trial registration:** ClinicalTrials.gov ID: NCT04452175. Registered June 29, 2020.

## INTRODUCTION

Smoking prevalence among people with mental disorders is about two to four times higher than in the general population (Tidey, 2015). People with schizophrenia spectrum disorders smokes more heavily and is more dependent on tobacco cigarette than those without mental illness (Zhang et al. 2013). Several potential contributing factors have been proposed, but the reason for the high prevalence of nicotine dependence and smoking in patients with schizophrenia spectrum disorders remains elusive.

As a result of high smoking rates, people with a mental health condition also have high rates of morbidity and mortality from smoking-related diseases compared with the general population (de Leon and Diaz, 2005; Callaghan et al., 2014; Olfson et al., 2015). Therefore, quitting smoking is particularly important for this group.

Progress in reducing smoking prevalence in people with mental health diagnoses has been very slow compared to the general population (Aubin et al., 2012; Le Cook 2014). Successful implementation of smoking cessation in people with schizophrenia is challenging. First, in people with schizophrenia the undesirable neurobiological and psychosocial consequences from stopping smoking are more pronounced and result in early relapse (Aubin et al., 2012). Second, they have lower appreciation of the health risks from smoking and are less interested in quitting compared to people without schizophrenia (Kelly et al., 2013). Third, the public health strategies used to curb smoking in the general population (i.e. tobacco taxation) appear to not be as effective among people with mental illness (Ashton et al., 2014). And fourth, smokers with schizophrenia spectrum are less likely to be offered smoking cessation treatment (Goldberg., 2010; Trainor et al., 2017) and when this happens, success rates are limited. Randomized clinical trials of smoking cessation treatment (varenicline, bupropion, nicotine replacement therapy) in schizophrenia have shown modest efficiency and only in the short term (Caponnetto et al, 2018).

Consequently, there is a pressing need for alternative and more efficient interventions to reduce or prevent morbidity and mortality in smokers with schizophrenia spectrum disorders. A realistic alternative is to encourage these people to eliminate or substantially reduce their exposure to tobacco smoke toxicants by switching to less harmful sources of nicotine delivery (such as electronic cigarettes, tobacco heated products or snus) and select the ones that give the greatest probability of abstaining from smoking (Polosa et al., 2013; O’Leary & Polosa 2020).

Electronic cigarettes (ECs) are continuing to gain popularity and acceptance by consumers worldwide for cigarette smoking substitution. Their design and efficiency in nicotine delivery have improved substantially since their market introduction more than 10 years ago. While the number of studies examining ECs effectiveness for smoking cessation and relapse prevention in the general population is now fairly substantial (Hartmann-Boyce et al., 2016; El Dib et al., 2017; Giovenco & Delnevo 2018), far fewer studies have been conducted with people with schizophrenia spectrum disorders (Caponnetto et al, 2018).

The first study to investigate the smoking cessation or reduction potential of e-cigarettes in people with schizophrenia, reported the efficiency of 12-week ad-libitum use of a rechargeable cigalike with 7.4 mg/ml nicotine cartridges in 14 people with schizophrenia, not motivated to stop smoking (Caponnetto et al. 2013). At 12 months follow up, 50% of smokers with schizophrenia had reduced their daily cigarettes consumption by at least 50%, and a further 14% had quit smoking completely with no increase in psychiatric symptoms (Caponnetto et al., 2013).

Subsequently, Pratt et al. (2016) studied 19 smokers with mental illness (10 with schizophrenia spectrum disorders, and 9 with bipolar disorder) who had no desire to quit. They were given a 4-week supply of a second-generation e-cigarette and were evaluated weekly for one month. Significant reduction in smoking (mean self-reported decline from 192 to 67 cigarettes per week confirmed by carbon monoxide expired air reduction) was reported.

Hickling et al. (2018) investigated the effectiveness and acceptability of a six-week supply of a disposable e-cigarette with 45 mg/ml nicotine to reduce smoking in 50 smokers with severe mental illness (including 42 with schizophrenia spectrum disorders) not motivated to quit. At the end of the six-week supply phase, 37% of participants had reduced their tobacco consumption and 7% had stopped smoking. Four weeks after the end of the 6-week supply, 26% of participants had reduced their tobacco consumption and 5% had quit combustible cigarettes. At final follow up (24 weeks), 25% of participants had reduced their tobacco consumption and 2% had quit to smoke combustible cigarettes.

In a more recent a single arm pilot study (Caponetto et al. 2018b), we have reported substantial changes in cigarette use behaviour amongst a group of 40 heavy smokers with a schizophrenia spectrum disorder diagnosis, who did not intend to reduce or quit smoking. Participants were invited to use a JUUL e-cigarette with 5% nicotine strenght for at least 12 weeks and then followed prospectively for up to 24 weeks. By the end of 12 weeks, 16 (40%) participants quit smoking and all were still using their e-cigarette. Overall, a sustained >50% reduction or smoking abstinence was reported in 37 (92.5%) of the study participants. We concluded that JUUL e-cigarette with 5% nicotine strength may have sufficient nicotine delivery and product appeal to help people with schizophrenia spectrum disorders to quit or reduce smoking and that further research with a larger sample and a comparator group would be needed.

In consideration of the preliminary evidence that ECs may be useful for smoking cessation and relapse prevention in people with schizophrenia spectrum disorders (Caponnetto et al, 2018) and that substantial quit rates were observed with high nicotine strenght JUUL e-cigarettes in smokers with schizophrenia (Caponetto et al. 2018b)., we hypothesized that switching smokers with a schizophrenia spectrum disorder diagnosis to JUUL e-cigarette with 5% nicotine strength could result in higher success rates compared to JUUL e-cigarette with 1.5% nicotine strength. Recent work indicates that nicotine PK of the JUUL e-cigarette with 5% nicotine strength (a device that utilizes a nicotine salt formulation) approximates the nicotine delivery of combustible cigarettes (Yingst et al., 2019; Hajek P et al., 2020; Maloney et al., 2020) and that the 5% nicotine strength product is far more efficient in delivering nicotine compared to the sister product with 1.5% nicotine strength (Talih et al., 2020). Both products are identical in their appearance, making them suitable for a double-blind study design.

Knowledge about the impact of ECs use on smoking habits of people with schizophrenia is limited and large randomized controlled trials are needed. Therefore, we have designed a 12-months prospective randomized trial of smokers with schizophrenia spectrum disorders to test the hypothesis that switching to ECs may accomplish high quit and reduction rates that persist long-term. Participants will be randomized to either high (JUUL 5% nicotine) or low nicotine strength devices (JUUL 1.5% nicotine) of identical appearance and assigned in a double-blind fashion. A much lower success rate is expected in the study group randomized to switching to low nicotine strength products compared to high nicotine strength. Besides effectiveness, tolerability, acceptability, and pattern of use of ECs will be compared between study groups.

In summary, the main objectives of the study will be to:

1. Quantify the proportion of **continuous quitters** among participants at 6-months in both arms of the study;
2. Quantify the proportion of **continuous reducers** among participants at 6-months in both arms of the study;
3. Quantify the proportion of continuous quitters among participants at 12-months in both arms of the study;
4. Quantify the proportion of continuous reducers among participants at 12-months in both arms of the study;
5. Compare continuous quit and reduction rates between study arms at 6- and 12-months;
6. Quantify **adverse events** throughout the whole duration of the interventional phase of the study in both arms;
7. Compare adverse events between study arms.

Additional objectives of the study will be to:

1. Measure **Subjective perceptions and experiences** of the two nicotine strenghts by the psychometrically validated modified Cigarette Evaluation Questionnaire - mCEQ (at 6-months);
2. Compare level of mCEQ between study arms (at 6-months);
3. Assess **pattern of products use** among participants throughout the whole duration of the study (both at intervention + follow-up phases) in both arms of the study;
4. Compare pattern of product use between study arms (both at intervention + follow-up phases).
5. Compare changes in **symptom severity** of patients with schizophrenia by Positive and Negative Syndrome Scale (PANSS) within and between both arms of the study (both at 6- and 12-months);
6. Compare changes in **exercise tolerance** by Chester Step Test within and between both arms of the study (only at 6-months);
7. Compare changes in **weight/BMI** within and between both arms of the study (both at 6- and 12-months);
8. Quantify **self-rated mental health** (SRMH) throughout the whole duration of the study (by means of a specifically designed APP) within and between both arms of the study (both at 6- and 12-months).

## METHODS

### TRIAL DESIGN

This will be a multicenter, 12-months prospective trial, utilizing a randomized, double-blind, 2-arm parallel, switching design to compare effectiveness, tolerability, acceptability, and pattern of use between high (JUUL 5% nicotine) and low nicotine strength devices (JUUL 1.5% nicotine) in adult smokers with schizophrenia spectrum disorders. The study will take place at international sites: UK (London), Italy (Milan, Rome and Catania) Russia (Ufa and St. Petersburg) and Ukraine (Odessa).

Participants will be randomized (1:1 ratio) to either high (JUUL 5% nicotine) or low nicotine strength devices (JUUL 1.5% nicotine) study arm (**Figure 1, 2)**. The two devices have identical appearance and will be assigned in a double-blind fashion. Study products will be provided for a total of 6-months (intervention phase); the intervention phase will be followed by a further 6-months observational period (follow-up phase) during which no products will be dispensed to participants. Throughout the follow-up phase and up to the final visit at 12-months, changes in smoking/vaping behavior and in their pattern of use will be tracked under naturalistic condition and compared between study groups. Any changes in symptom severity related to schizophrenia spectrum disorders within and between both arms of the study will be monitored. The intervention phase of the study will consist of a total of nine visits (including screening). The follow-up phase will consist of three additional visits (two telephone contacts and one final face to face visit).

**Figure 1.**
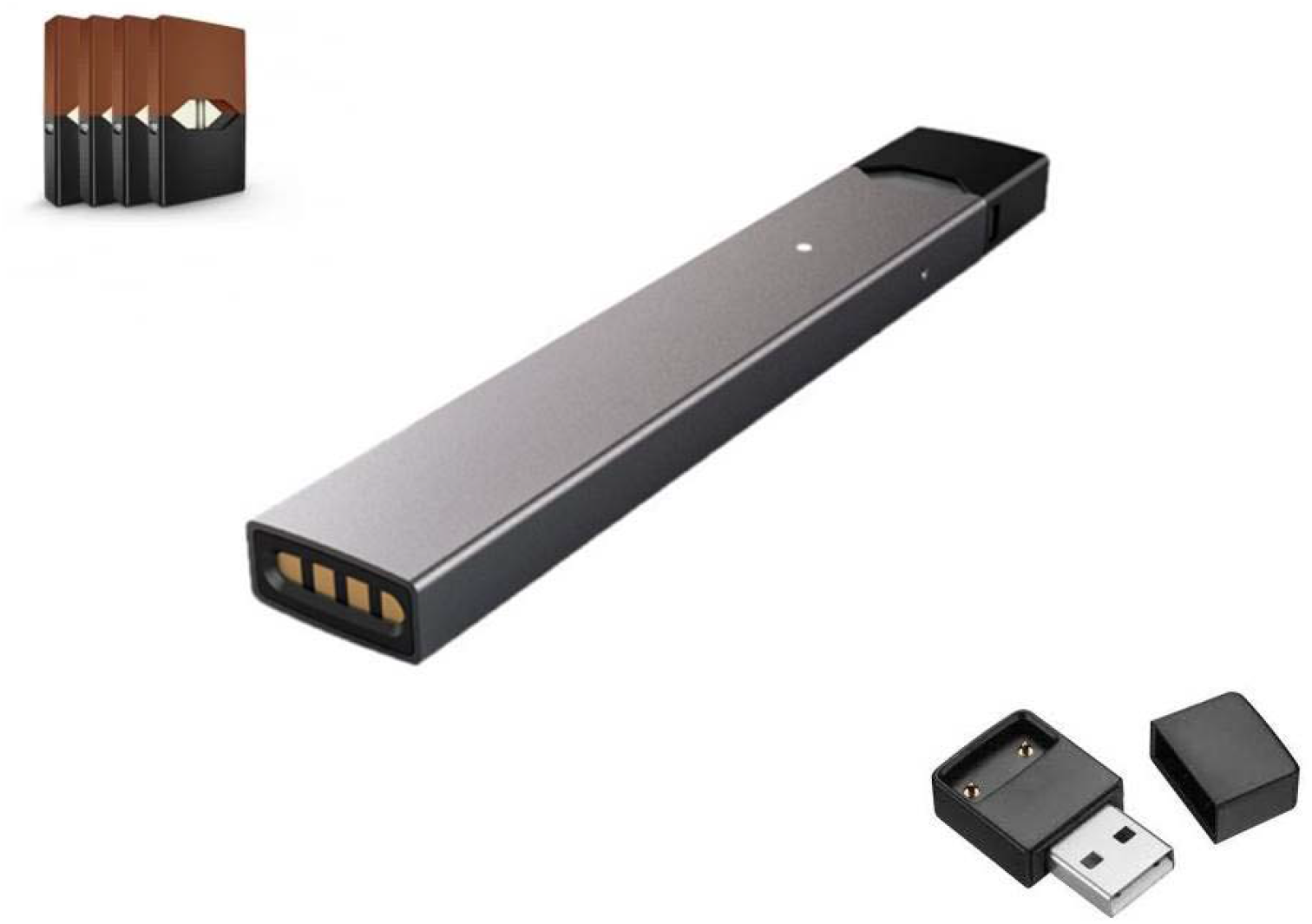

**Figure 2.**
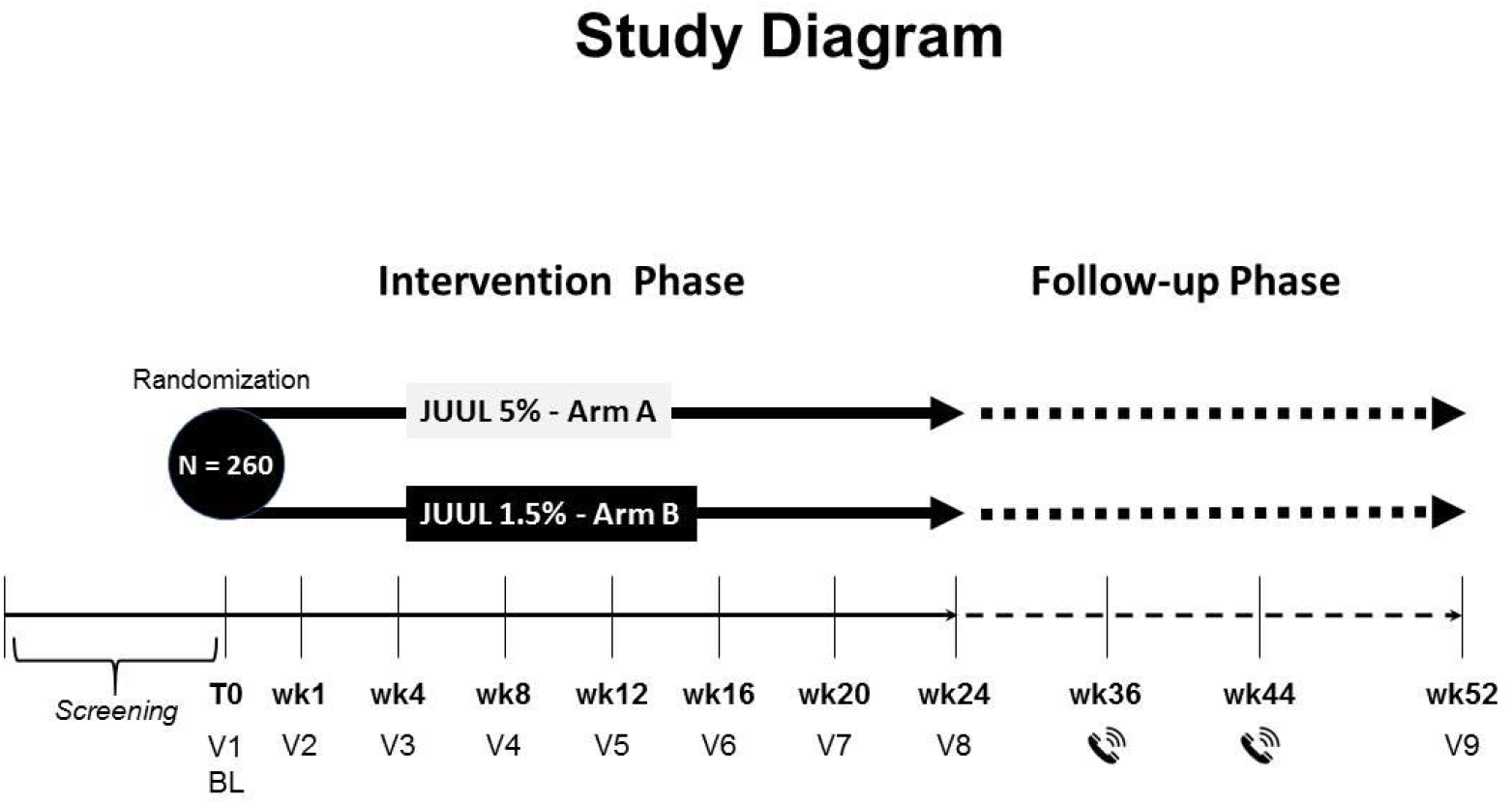

The design of the trial follows the rules set by the Consolidated Standards of Reporting Trials (CONSORT) guidelines.

### STUDY PARTICIPANTS

Adult non-hospitalized daily smokers (>10 cigarettes per day), meeting criteria for a schizophrenia spectrum disorder diagnosis without evidence of recent exacerbation of illness (defined as “no relapse to hospitalization within the last 3 months and no change antipsychotic treatment within the last month” - Mendrek, 2012) will be recruited. The diagnosis of schizophrenia spectrum disorder will be based on the criteria from the 5th edition of the Diagnostic and Statistical Manual of Mental Disorders (American Psychiatric Association, 2013).

At screening and prior to enrolment, all patients will be offered a locally available free smoking cessation program as per local guidelines. Those who express the intention of booking for the cessation program together with those who, at screening, are planning to quit smoking in the next month, will not be recruited in the study. Patients taking part in the study will be informed that they are free to quit smoking and withdraw from the study at any time. Participants will be encouraged to quit smoking at every contact timepoint throughout the whole study. Any participant who decides to quit smoking will be directed to local cessation services.

Volunteers will be recruited from outpatient mental health clinics/or psychiatric practices. According to sample size calculation (see below) we aim at recruiting 258 regular smokers with schizophrenia. It is estimated that the enrolment period will last about 6 months.

### CONSENT

At each site the Principal Investigator (PI) will retain overall responsibility for the conduct of research; this includes the taking of informed consent of participants at their site. They must ensure that any person delegated responsibility to participate in the informed consent process is duly authorised, trained and competent to participate according to the ethically approved protocol, Principles of Good Clinical Practice (GCP) and Declaration of Helsinki.

At the screening visit potential participants will be provided by clinic staff with a verbal outline of the project, accompanied by a written patient information sheet to take home and read at leisure. Once eligibility has been established and screening procedures completed, participants will be invited to the baseline visit.

At the baseline check, after confirmation of eligibility and before randomization, all smokers will be reminded of the risks associated with smoking and will be offered a free smoking cessation program according to standard local guidelines and depending on the local availability of antismoking services. Those who decline the invitation will be eligible for recruitment into the study.

These potential participants will be asked whether they wish to enter the study. The PI or a delegated and suitably trained member of clinical staff will provide further explanation of the study and practical details; and give an opportunity for questions to be answered; before obtaining participants’ written informed consent.

Participation is voluntary, and participants may refuse to participate, or to withdraw from the study at any stage without having to give an explanation, and with no impact on their treatment.

### ELIGIBILITY CRITERIA

#### Inclusion criteria

Participants will be required to satisfy all of the following criteria unless otherwise stated (these will be confirmed at screening and at baseline visit):

- Adult (≥ 21yrs)
- Regular smoking (>10 cigarettes a day; for at least one year)
- Exhaled breath CO (eCO) level > 7 ppm
- Not currently attempting to quit smoking or wishing to do so in the next 30 days; this will be verified at screening by the answer ‘‘NO’’ to the question ‘‘Do you intend to quit in the next 30 days?’’
- Schizophrenia spectrum disorder diagnosis (schizophrenia, delusional disorder, schizoaffective disorder, personality disorder, schizoid personality disorder, etc) by DSM-V criteria
- Understand and provide informed consent
- Able to comply with all study procedures

#### Exclusion criteria

Participants will be excluded based on the following criteria (these will be confirmed at screening and at baseline visit):

- Institutionalized patients
- Acute decompensation of Schizophrenia spectrum disorder symptoms within the past month
- Change in antipsychotic treatment within the past month
- No recent history of hospitalization for any serious medical condition within 3 months prior to screening, as determined by the investigator.
- Myocardial infarction or angina pectoris within 3 months prior to screening, as determined by the investigator.
- Current poorly controlled asthma or COPD
- Pregnancy, planned pregnancy or breastfeeding. Any female participant who becomes pregnant during this study will be withdrawn.
- Participants who have a significant history of alcoholism or drug/chemical abuse within 12 months prior to screening, as determined by the investigator.
- Accepting to take part in a smoking cessation program
- Participants who regularly use any recreational nicotine (e.g. e-cigarettes,) or tobacco product (e.g. tobacco heated products, oral smokeless) other than their own cigarettes within 30 days of screening.
- Participants who have used smoking cessation therapies (e.g varenecline, buproprion, or NRT) within 30 days of screening.
- Participants who are still participating in another clinical study (e.g. attending follow-up visits) or who have recently participated in a clinical study involving administration of an investigational drug (new chemical entity) within the past 3 months.
- Participants who have, or who have a history of, any clinically-significant neurological, gastrointestinal, renal, hepatic, cardiovascular, psychiatric, respiratory, metabolic, endocrine, haematological or other major disorder that, in the opinion of the investigator or their appropriately qualified designee, would jeopardise the safety of the participant or impact on the validity of the study results.

### RANDOMIZATION

Participants who are eligible and consent to take part will be randomized (1:1 ratio) to receive 6-month of either high (JUUL 5% nicotine; Study Arm A) or low nicotine strength devices (JUUL 1.5% nicotine; Study Arm B) (**Figure 1)**. The intended number of participants in each arm of the study is 129 (258 in total).

The randomization sequence will be computer generated and provided to clinical sites via a web-based application set up by the CRO. The staff randomizing the participant will access the web-based application when the participant is with them, entering their participant identification number, date of birth and initials into the program. The allocation will be immediately provided by the program/software.

Before randomization, all participants will be reminded of the risks associated with smoking and will be offered a free smoking cessation program according to standard local guidelines and depending on the local availability of antismoking services. Those who decline the invitation will be eligible for randomization.

After randomization, and throughout the study, participants will be explicitly told that they should be quit smoking and they will receive brief smoking cessation adviceat each contact. Participants are free to withdraw from the study at any time.

### STUDY PRODUCTS

JUUL e-cigarette is a popular vapor product consisting of two main parts: a battery body and a pre-filled pod that connects to the battery body. The pod is pre-filled with a 0.7 ml proprietary e-liquid formulation that combines glycerol, propylene glycol, flavourings, nicotine and benzoic acid. JUUL is breath actuated and has no user modifiable settings. The battery (200 mAh) is not removable, and it is rechargeable via an USB port. The device is also equipped with a unique temperature regulation feature that has been demonstrated to maintain coil and wick temperatures below 300 C under a range of operating conditions. This feature helps avoiding the “dry puff” phenomenon and the excess generation of carbonyls in the aerosol emissions that may be associated with it.

JUUL e-cigarette with tobacco flavoured 5% nicotine strength pods has been chosen for the following reasons:

1. Its nicotine delivery is comparable to that of a traditional cigarette (Yingst et al., 2019; Hajek P et al., 2020; Maloney et al., 2020);
2. It is very easy to use (the pod system design does not require manual re-filling);
3. High success rates in smokers with schizophrenia spectrum disorders have been shown with this product (Caponnetto et al., 2018b).

The reference group will use tobacco flavoured 1.5% nicotine strength pods. We restricted flavor options to regular tobacco flavor to most closely match usual cigarette brand flavor profile and reduce unwanted variance in product. Tobacco flavoured 5% and 1.5% nicotine pods are identical in appearance, and therefore suitable for double-blinding.

### BLINDING

Blinding is ensured by the identical appearance of the device and pods. Participants and research staff conducting study sessions will be blinded to the nicotine strength in the pods. In order to mask 5% vs 1.5% tobacco flavour pods, a designated member of hospital pharmacy staff - who is not involved in the study - will remove the labeling from the pod blister. Unlabeled pod blisters will be repackaged in a coded zip-lock plastic bag. Blinded data will be seen and analysed by the Trial Statistician for the purposes of the Data Monitoring and Ethics Committee (DMEC) meetings. All other trial staff who have access to outcome data will remain blinded until prespecified data analyses has be completed. Prespecified data analyses will be conducted blind to treatment allocation.

### STUDY VISITS

The study will consist of a 6-month intervention phase and a 6-month follow-up phase (Figure 1). A total of nine visits (including screening) are scheduled during the intervention phase; a screening visit, a baseline (BL) visit and seven study visits (at weeks 1, 4, 8, 12, 16, 20, and 24). The participants will attend their study visits in an outpatient clinic setting at approximately the same time of day. With the exception of the baseline (BL) study day, most visits will take approximately 30 to 45 minutes to complete. Three additional visits are scheduled for the follow-up phase - two managed via telephone contacts (TC) at weeks 36 and 44, and one final onsite visit at weeks 52. The **study schedule in table 1** describe the procedures to be completed at each visit.

**Table 1.**
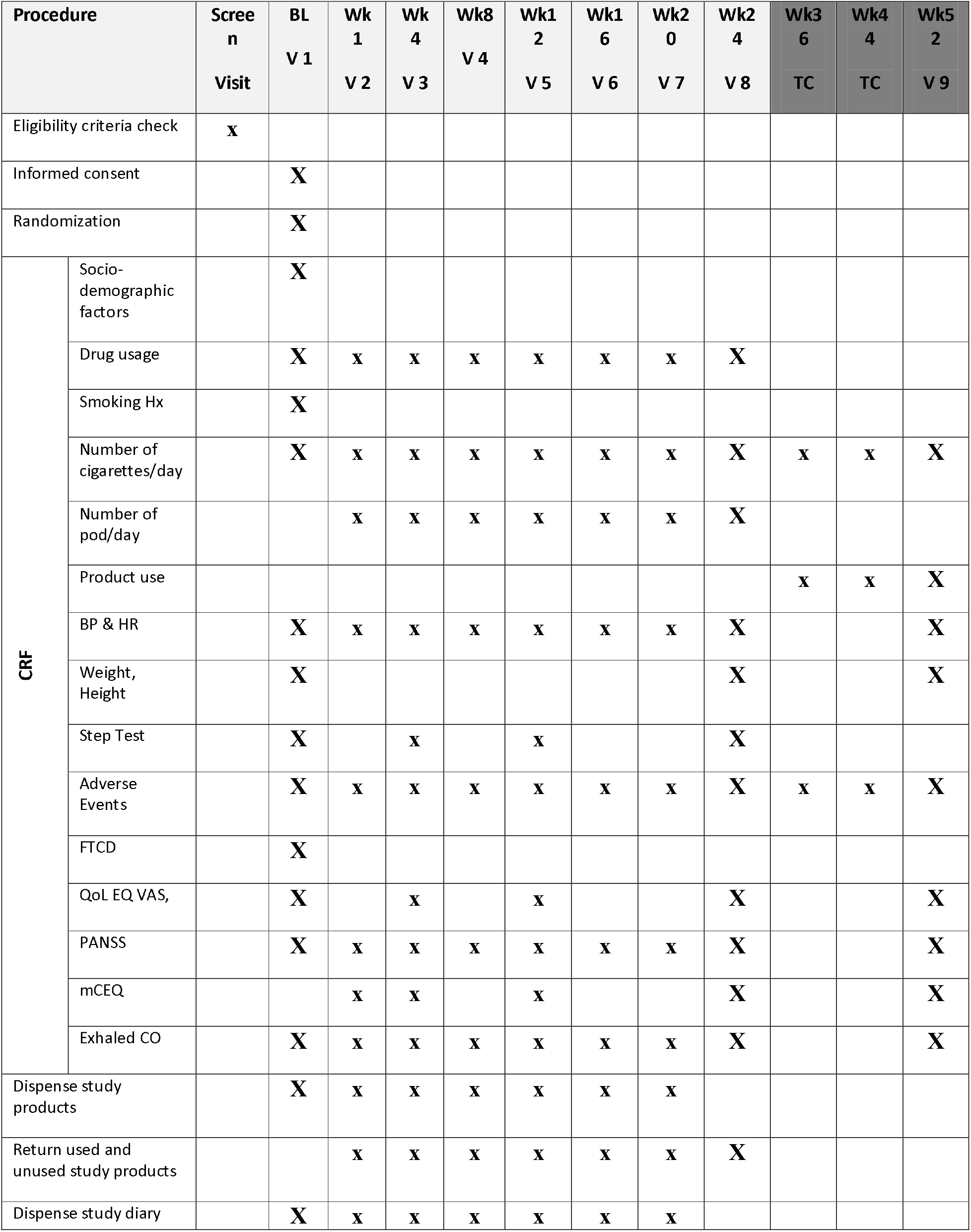
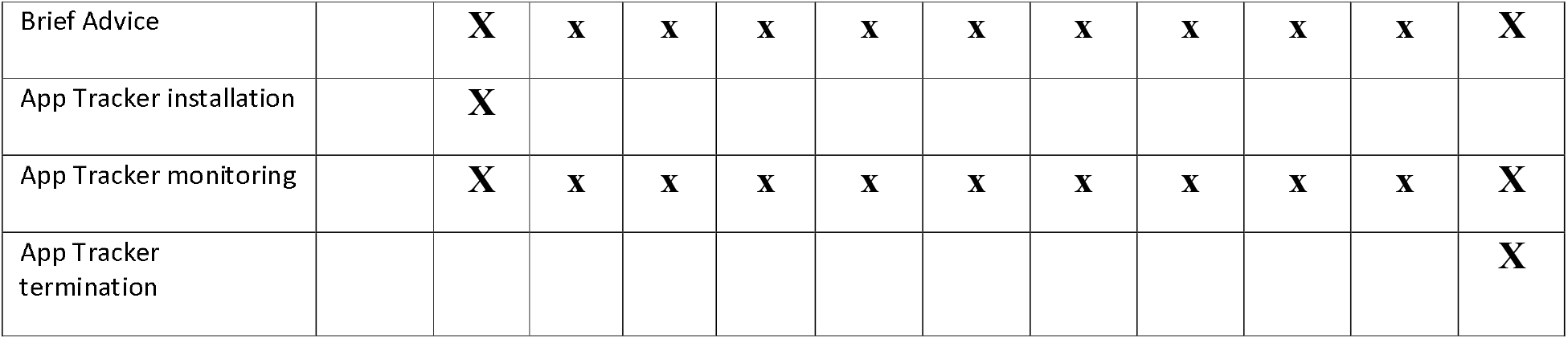

#### Screening Visit (Visit 0)

Potential participants will attend a screening visit to assess eligibility criteria. The objective of the research will be discussed by a clinical site staff member and patient will be asked to take the time to read and review the study-specific Patient Information Sheet. All patients will be offered smoking cessation program as per local guidelines, and if they choose to accept the program they will be directed to local smoking cessation services and will not take part in the study. Eligibility criteria check includes socio-demographic data (sex, age, and ethnicity), medical history (including medication use), and smoking history. Assessments conducted at the Screening Visit are presented in Table 1. Eligible patients will be invited to attend a baseline visit.

#### Baseline Visit (Visit 1)

Baseline visit will be carried out within 4 weeks of the screening visit. Prior to check-in, participants will refrain from smoking for at least 30 min. Intention to quit will be again emphasised. Patients expressing the intention of quitting in the next 30 days will be referred to the local smoking cessation services and will not be recruited in the study.

All other participants will sign the informed consent form and undergo the following assessments prior to enrolment/randomization (Table 1):

- Review of inclusion/exclusion criteria
- Completion of CRF
  - socio-demographic data
  - medical and medication hx
  - AEs
  - smoking hx
  - number of cigarettes consumed on a daily basis
  - Fagerstrom questionnaire for Cigarette Dependence (FTCD)
- Review of pregnancy status

Following enrolment, participants will undergo the following baseline assessments (Table 1):

- Vital signs (BP, HR)
- Height and body weight
- Positive and Negative Syndrome Scale (PANSS)
- Carbon monoxide breath tests (eCO)
- QoL (EQ VAS)
- Step Test

Following baseline assessments, participants will be randomized to receive 6-month of either high (JUUL 5% nicotine; Study Arm A) or low nicotine strength pods (JUUL 1.5% nicotine; Study Arm B) (**Figure 1)**.

After randomization, participants will be given a JUUL device and asked to trial and familiarize with the nicotine strength pod to which they were assigned. Participants will be explicitly told that they should be quit smoking and they will receive brief smoking cessation advice at each contact. The brief advice has been specifically designed to help smokers with mental health problems to quit or reduce smoking (Robson & Pots, 2014; Robson & McEwen, 2018).

A prospective evaluation of cigarette consumption is an important feature of this type of studies. Self-reported cigarette smoking and vaping product use can be tracked throughout the study via APP technology. The App tracker prompts a few simple queries on a daily basis, and participants have the possibility to record this information. The importance of product adherence and compliance to smoking abstinence throughout the study will be explained to participants and a specifically designed tracker APP will be installed in their smartphones to record number of cigarette smoked per day, and pod usage. Non-compliance will be documented and analysed. The App will also keep track of symptoms/AEs (related to nicotine abuse, nicotine withdrawal, antipsychotic drugs toxicity, and decompensation of schizophrenia).

Besides the tracker APP, paper based study diaries will be also used to record cigarettes and pods consumption as well as symptoms/AEsAEs.

Prior to check-out, participants will receive a 1-week study diary, a JUUL device and 1-week supply of either 5% or 1.5% nicotine pods depending on the study-arm allocation. They will receive a number of pods per day corresponding to the number of cigarette smoked at baseline (e.g. a smoker of 10-20 cig/day will receive 1pod/day, a smoker of 21-40 cig/day will receive 2pods/day, a smoker of 41-60 cig/day will receive 3pods/day).

Participants will be invited to return to the next programmed study visit (study visit 2, at week 1), to complete the programmed study assessments and to obtain their next supply of nicotine pods.

#### Week-1 Visit (Visit 2)

Visit 2 will be carried out within one week of visit 1. Prior to check-in, participants will refrain from smoking for at least 30 min. Completed study diaries and used and unused study products will be returned by the participants to the site investigator.

Participants will receive brief smoking cessation advice and undergo the following study assessments (Table 1):

- Review of medication use
- Symptoms/AEs
- Number of cigarettes consumed on a daily basis
- Pod usage
- Vital signs (BP, HR)
- Positive and Negative Syndrome Scale (PANSS)
- Carbon monoxide breath tests (eCO)
- Modified Cigarette Evaluation Questionnaire (mCEQ)
- App tracking data for the study period will be downloaded and verified

Prior to check-out, participants will receive a 3-week study diary, and 3-week supply of either 5% or 1.5% nicotine pods depending on the study-arm allocation.

Participants will be invited to return to the next programmed study visit (study visit 3, at week 4), to complete the programmed study assessments and to obtain their next supply of nicotine pods.

#### Week-4 Visit (Visit 3)

Visit 3 will be carried out within three weeks of visit 2. Prior to check-in, participants will refrain from smoking for at least 30 min. Completed study diaries and used and unused study products will be returned by the participants to the site investigator.

Participants will receive brief smoking cessation advice and undergo the following study assessments (Table 1):

- Review of medication use
- Symptoms/AEs
- Number of cigarettes consumed on a daily basis
- Pod usage
- Vital signs (BP, HR)
- Chester Step Test
- QoL (EQ VAS)
- Positive and Negative Syndrome Scale (PANSS)
- Carbon monoxide breath tests (eCO)
- Modified Cigarette Evaluation Questionnaire (mCEQ)
- App tracking data for the study period will be downloaded and verified

Prior to check-out, participants will receive a 4-week study diary, and 4-week supply of either 5% or 1.5% nicotine pods depending on the study-arm allocation.

Participants will be invited to return to the next programmed study visit (study visit 4, at week 8), to complete the programmed study assessments and to obtain their next supply of nicotine pods.

#### Week-8 Visit (Visit 4)

Visit 4 will be carried out within four weeks of visit 3. Prior to check-in, participants will refrain from smoking for at least 30 min. Completed study diaries and used and unused study products will be returned by the participants to the site investigator.

Participants will receive brief smoking cessation advice and undergo the following study assessments (Table 1):

- Review of medication use
- Symptoms/AEs
- Number of cigarettes consumed on a daily basis
- Pod usage
- Vital signs (BP, HR)
- Positive and Negative Syndrome Scale (PANSS)
- Carbon monoxide breath tests (eCO)
- App tracking data for the study period will be downloaded and verified

Prior to check-out, participants will receive a 4-week study diary, and 4-week supply of either 5% or 1.5% nicotine pods depending on the study-arm allocation.

Participants will be invited to return to the next programmed study visit (study visit 5, at week 12), to complete the programmed study assessments and to obtain their next supply of nicotine pods.

#### Week-12 Visit (Visit 5)

Visit 5 will be carried out within four weeks of visit 4. Prior to check-in, participants will refrain from smoking for at least 30 min. Completed study diaries and used and unused study products will be returned by the participants to the site investigator.

Participants will receive brief smoking cessation advice and undergo the following study assessments (Table 1):

- Review of medication use
- Symptoms/AEs
- Number of cigarettes consumed on a daily basis
- Pod usage
- Vital signs (BP, HR)
- Chester Step Test
- QoL (EQ VAS)
- Positive and Negative Syndrome Scale (PANSS)
- Carbon monoxide breath tests (eCO)
- Modified Cigarette Evaluation Questionnaire (mCEQ)
- App tracking data for the study period will be downloaded and verified

Prior to check-out, participants will receive a 4-week study diary, and 4-week supply of either 5% or 1.5% nicotine pods depending on the study-arm allocation.

Participants will be invited to return to the next programmed study visit (study visit 6, at week 16), to complete the programmed study assessments and to obtain their next supply of nicotine pods.

#### Week-16 Visit (Visit 6)

Visit 6 will be carried out within four weeks of visit 4. Prior to check-in, participants will refrain from smoking for at least 30 min. Completed study diaries and used and unused study products will be returned by the participants to the site investigator.

Participants will receive brief smoking cessation advice and undergo the following study assessments (Table 1):

- Review of medication use
- Symptoms/AEs
- Number of cigarettes consumed on a daily basis
- Pod usage
- Vital signs (BP, HR)
- Positive and Negative Syndrome Scale (PANSS)
- Carbon monoxide breath tests (eCO)
- App tracking data for the study period will be downloaded and verified

Prior to check-out, participants will receive a 4-week study diary, and 4-week supply of either 5% or 1.5% nicotine pods depending on the study-arm allocation.

Participants will be invited to return to the next programmed study visit (study visit 7, at week 20), to complete the programmed study assessments and to obtain their next supply of nicotine pods.

#### Week-20 Visit (Visit 7)

Visit 7 will be carried out within four weeks of visit 4. Prior to check-in, participants will refrain from smoking for at least 30 min. Completed study diaries and used and unused study products will be returned by the participants to the site investigator.

Participants will receive brief smoking cessation advice and undergo the following study assessments (Table 1):

- Review of medication use
- Symptoms/AEs
- Number of cigarettes consumed on a daily basis
- Pod usage
- Vital signs (BP, HR)
- Positive and Negative Syndrome Scale (PANSS)
- Carbon monoxide breath tests (eCO)
- App tracking data for the study period will be downloaded and verified

Prior to check-out, participants will receive a 4-week study diary, and 4-week supply of either 5% or 1.5% nicotine pods depending on the study-arm allocation.

Participants will be invited to return to the next programmed study visit (study visit 8, at week 24), to complete the programmed study assessments and to obtain their next supply of nicotine pods.

#### Week-24 Visit (Visit 8)

Visit 8 will be carried out within three weeks of visit 2. Prior to check-in, participants will refrain from smoking for at least 30 min. Completed study diaries, JUUL device and used and unused study products will be returned by the participants to the site investigator.

Participants will receive brief smoking cessation advice and undergo the following study assessments (Table 1):

- Review of medication use
- Symptoms/AEs
- Number of cigarettes consumed on a daily basis
- Pod usage
- Vital signs (BP, HR)
- Weight, height
- Chester Step Test
- QoL (EQ VAS)
- Positive and Negative Syndrome Scale (PANSS)
- Carbon monoxide breath tests (eCO)
- Modified Cigarette Evaluation Questionnaire (mCEQ)
- App tracking data for the study period will be downloaded and verified

This study visit marks the end of the intervention phase and the beginning of the follow up phase. At this point no more study products will be dispensed by site investigators. Prior to check-out, participants expressing the intention of quitting will be referred to the local smoking cessation services. All the others will receive a leaflet summarizing local retail outlets and secure websites where to buy vaping products in case they wish to do so.

Participants will be invited to return to a final study visit (study visit 9, at week 52), to complete the programmed study assessments and reminded that two separate telephone contacts (TC) will be scheduled at weeks 36 and 44 to review product usage and smoking behavior under naturalistic condition of use.

#### Week-36 and Week-44 Telephone Contact Visits

These two telephone contacts (TC) will be carried out to review product usage and smoking behavior under naturalistic condition of use (over the last 12-weeks). At each TC, information on any symptoms/AEs will be also collected and a brief smoking cessation advice will be offered over the phone.

The following information (over the last 12-weeks) will be collected:

- Symptoms/AEs
- Number of cigarettes consumed on a daily basis
- Vaping (or any other nicotine) product usage (if any)
- App tracking data for the study period will be downloaded and verified

Participants will receive brief smoking cessation advice over the phone and will be reminded of the final study visit.

#### Week-52 Visit (Visit 9)

Visit 9 will be carried out within 6-months of visit 8. Prior to check-in, participants will refrain from smoking for at least 30 min. Participants will receive brief smoking cessation advice and undergo the following study assessments (Table 1):

- Symptoms/AEs
- Number of cigarettes consumed on a daily basis
- Vaping (or any other nicotine) product usage (if any)
- Vital signs (BP, HR)
- Weight, height
- QoL (EQ VAS)
- Positive and Negative Syndrome Scale (PANSS)
- Carbon monoxide breath tests (eCO)
- Modified Cigarette Evaluation Questionnaire (mCEQ)
- App tracking data for the study period will be downloaded and verified
- App tracking will be disinstalled

This study visit marks the end of the follow up phase and of the study.

### STUDY OUTCOMES MEASURES

The primary outcome measure for the study will be continuous smoking abstinence at 6-month; defined as self-reported continuous smoking abstinence at 6-month from the previous visit, biochemically verified by exhaled CO measurements of ≤ 7 ppm.

Secondary outcome measures will include quantitative within- and between-group variations from baseline of the following study endpoints:

- Proportion of continuous smoking abstinence at 12-month
- Proportion of continuous smoking reduction at 6-month
- Proiportion of continuous smoking reduction at 12-month
- Proportion of AEs
- Absolute change in PANSS
- Absolute change in mCEQ
- Absolute change in Chester Step Test-derived values
- Change in App-derived endpoints (self-rated mental health -SRMH).

Safety assessments will be conducted at each study visit. The safety analysis set (SAF) will include all participants who received any randomised study products, regardless of their achievement of substantial smoking cessation. This analysis set will be used to summarize safety information.

### SAFETY AND TOLERABILITY

All Adverse Events (AEs) and Serious Adverse Events (SAEs) with suspected causal relationships to study (e.g. related to mental health, tobacco cigarette smoking, e-cigarette use, and to withdrawal from nicotine or nicotine overdosing), will be noted and recorded during the whole duration of the study. AEs and SAEs will be recorded from baseline onwards and at each subsequent study visit on the adverse event page of the eCRF. Signs or symptoms will be elicited at each visit by open questioning, such as “How have you been feeling since your last visit?”. Subject will also be encouraged to spontaneously report AEs occurring at any other time during the study via the dedicated mobile APP or the communication channels provided for the study. The study investigator must pursue and obtain information adequately both to determine the outcome of the AE and to assess whether it meets the criteria for classification as a SAE requiring immediate notification to the competent authority. Sufficient information should be obtained to assess causality. Follow-up of the (S) AE has to be done until its resolution longest until the end of the study.

### SAMPLE SIZE

This will be a large double blind RCT that will use the JUUL e-cigarette with different nicotine levels 5% and 1,5% in smokers with schizophrenia spectrum disorders, the first of its kind. The 24 week continuous abstinence rates used in estimating this sample size were extracted from one single arm pilot study. In this study (Caponnetto et al., 2013), the researchers proposed e-cigarettes to 14 smokers with schizophrenia spectrum disorders, not motivated to quit, and participants experimented one nicotine strengths (2.4%) and finally showed a quit rate of 14% at 6 months.

A sample size of 258 participants (129 per each arm) will be aimed for in this RCT. A 14% quit rate is expected in low nicotine level arm while, by doubling the nicotine level (5%), a doubling of quit rate (28%) is assumed in 5% high nicotine level arm. The sample size was calculated in line with the following parameters: 14% quit rate in 1.5% low nicotine level arm and 28% quit rate in 5% high nicotine level arm; significance level (alpha) = 0.05; power (1-ß) = 0.80. The smokers will be randomized into the two arms of our study protocol (i.e., 129 smokers for each arm).

### STATISTICAL METHODS

Continuous variables will be described as mean and standard deviation (for normally distributed variables), or median and interquartile range (for not normally distributed variables). Categorical variables will be described with percentages and absolute frequencies.

The differences in continuous variables between the two groups will be evaluated by the Kruskal-Wallis test, The differences between the two groups for normally distributed data will be evaluated by one-way analysis of variance (ANOVA). The normality of data distribution will be evaluated by means of the Kolmogorov-Smirnov test. Any correlation between the variables under evaluation will be assessed by Spearman r correlation. To analyze differences in frequency distribution of categorical variables we will use the Chi-square test with the Yates correction or the Fisher exact test. All statistical tests are two-tailed and are considered to be statistically significant at P < 0.05. The consistency of effects for pre-specified subgroups will be assessed using tests for heterogeneity. Subgroups will be based on age, sex, education, level of nicotine dependence.

Smokers who leave the study before its completion due to lack of efficacy or poor tolerability of the product under investigation, will carry out the early termination visit and will be defined as continuous smokers. AEs, symptoms thought to be related to tobacco smoking and e-cigarette use, and to withdrawal from nicotine, will be annotated at each subsequent study visit to BL on the AE page of the study diary. The safety analysis set (SAF) will include all participants who received any randomised treatment, regardless of their achievement of substantial smoking cessation. This analysis set will be used to summarize safety information. Participants will be analysed according to actual treatment received. Descriptive statistics will summarize the number and percentage of subjects experiencing adverse events by study group. Continuous data will be expressed as mean (±SD) and median (and interquartile range [IQR]). Within-group (from baseline) and between-group differences will be evaluated by means of statistical tests, for paired and unpaired continuous variables, as appropriate. Significance of differences in frequency distribution of categorical variables will be tested by χ2 test, at an alpha level of 0.05. The number and the percentage of subjects experiencing AEs, adverse reactions, serious AEs and AEs leading to study withdrawal will be summarized by treatment group. AEs will also be summarized by system organ class and preferred term using the MedDRA dictionary. Statistical analysis will be performed with SPSS 23.0 (Statistical Package for Social Sciences Program, IBM).

A logistic regression model will be fitted to binary endpoints and will include treatment and center as independent variables. Subjects who discontinue the study are assumed to be smokers for the remainder of the study. In responder rates, those subjects will be represented in the denominator but not in the numerator. For the endpoints that are continuous, a linear model including treatment and center will be used as the underlying model.

### WITHDRAWAL CRITERIA

Patients may be withdrawn from the study prematurely for the following reasons:

1. Participant experiences a severe adverse event (SAE). The appropriate SAE electronic Case Report Form (eCRF) page must be completed.
2. If any deviations occur during the conduct of the study, which cannot be corrected. All protocol deviations will be fully documented and considered for their effect on study objectives. Deviations that could lead to participant discontinuation from the study include:
  - deviations which could affect participant’s safety (e.g. illness requiring treatment[s]) which in the clinical judgement of the investigator might invalidate the study by interfering with the allocated test product or the willingness of the participant to comply with the study activities.
  - deviations involving the use of any nicotine/tobacco products other than the intended conventional cigarettes (in Arm A) or (in Arm B).
3. If the participant is uncooperative, including non-attendance. In these cases, efforts should have been made by the investigator to ascertain the reason and to ensure the participant’s attendance as soon as possible.
4. Participant’s personal request: the participant could decide, at any moment of the study, to stop his/her participation.
5. Female participant becoming pregnant.

### TRIAL MEASUREMENTS

#### Chester Step Test

This is a simple validated test for assessing participants’ arerobic capacity (i.e. exercise tolererance). The participant has to perform sequential climbs and descents from the step following the rhythm dictated by the metronome. Test will be carried out at the times indicated in the Study Schedule.

#### QoL Scale

A validated scale for measuring self perceptions of the impact of quality of life will be completed by the participant - EQ VAS (Rabin & De Charro, 2001). The EQ VAS is a valid and reliable measure in many disease areas (Wailoo et al.,2010) and it is the most widely used generic patient reported outcome questionnaire (Devlin & Brooks, 2017). Questionnaires will be provided in the local language.

#### Fagerström Test for Cigarette Dependence

Cigarette dependence will be assessed via a FCTD questionnaire in its revised version (Fagerstrom, 2012). The questionnaire consists of 6 questions which will be answered by the participant himself/herself. The scores obtained on the test permit the classification of nicotine dependence into 3 levels: Mild (0-3 points), moderate (4-6 points), and severe (7-10 points). Questionnaires will be provided in the local language.

#### Exhaled Carbon Monoxide

Participants’ smoking status will be objectively verified by a CO breath test (exhaled CO >7 ppm). Levels of carbon monoxide in exhaled breath (eCO) will be measured using a portable CO monitor (MicroCO, Vyaire, or similar device). Participants will not be allowed to vape or smoke within 30 minutes prior to eCO level measurements. Levels will be measured at the times indicated in the Study Schedule.

#### Product-use and self rated mental health (SRMH) Count/APP tracker

The number of conventional cigarettes smoked or products used will be self-reported and recorded in the e-CRF daily for the first 30 days of the study, and daily for 3 to 5 days prior to each clinic visit for the remainder of the study. In addition, the APP will track cigarette and e-cigarette use daily and self rated mental health (SRMH) throughout the whole duration of the study.

#### Positive and Negative Syndrome Scale (PANSS)

The Positive and Negative Syndrome Scale for Schizophrenia (PANSS) will be used for the assessment of schizophrenia symptom (Kay et al, 1987). It is used utilized frequently in clinical and research settings and its reliability and validity has been shown to be consistent in multiple cross-cultural settings. Questionnaires will be provided in the local language.

#### Modified Cigarette Evaluation Questionnaire (mCEQ)

This 11-items self-administered questionnaire will assesse the degree to which subjects experience the reinforcing effects of smoking or vaping (Cappelleri et al., 2007). In particular, evaluates level of enjoyment of respiratory tract sensations, craving reduction for cigarettes, psychological reward, aversions, and satisfaction. mCEQ use three multi-item domains (subscales) and two single items: “Smoking Satisfaction” (items 1, 2, plus item 12); “Psychological Reward” (items 4-8); “Aversion” (items 9,10); “Enjoyment of Respiratory Tract Sensations” (item 3); and “Craving Reduction” (item 11). Scores for each subscale are calculated as the average of its individual item responses. Questionnaires will be provided in the local language.

### CONCOMITANT MEDICATION

If any medication is required during this study, the name, strength, frequency of dosing and reason for its use will be documented in the participant’s eCRF by the Investigator. No illicit drugs will be taken.

## RESULTS

Recruitment of participants will start in Dec 2020 and enrolment is expected to be completed in May 2021. Final results will be reported in 2022.

## DISCUSSION

There is a pressing need for alternative and more efficient interventions to reduce or prevent morbidity and mortality in smokers with schizophrenia spectrum disorders. A realistic alternative is to encourage these people to eliminate or substantially reduce their smoking by switching to less harmful sources of nicotine delivery such as electronic cigarettes or any other source that gives the greatest probability of abstaining from cigarettes (Polosa et al., 2013; O’Leary & Polosa 2020). Little is known about the impact of e-cigarette use on people with schizophrenia spectrum disorders who smoke (Caponnetto et al, 2018). GENESIS will be the first study determining the overall health impact of e-cigarettes in people with schizophrenia spectrum disorders. Undoubtedly, it is desirable for these patients to avoid consumption of any tobacco-related inhalation products, but in order for Governments, health professionals and caregivers to provide guidance about cigarette substitution, robust evidence-based information is needed.

We designed this large multicenter RCT to gather such evidence. In particular, we will be testing the hypothesis that switching to e-cigarettes may result in substantial smoking abstinence in people with schizophrenia spectrum disorder who smoke. In addition, the GENESIS trial will provide the opportunity to assess the impact of e-cigarettes switching on several mental health parameters (including Positive and Negative Syndrome Scale - PANSS and self-rated mental health - SRMH) throughout the whole duration of this 1-year study. Long-term evaluation of these parameters is important to fully understand the risk/benefit profile of smoking substitution by vaping on mental health. The decision for a switching study design in GENESIS has been guided by the notion that e-cigarettes are widely used as substitutes for tobacco cigarettes (Caponnetto et al., 2012; Caponnetto et al., 2020, O’Leary R & Polosa 2020). The length of the trial is based on the consideration that changes in the primary endpoint could be reasonably observed as early as 6-months. It is however possible that a much longer follow-up period could be necessary to firmly establish findings consistency over time, hence follow up was extended to 12-months. The RCT study design will provide a robust answer to determine success rates and health impact of e-cigarettes in people with schizophrenia spectrum disorder. Clearly, randomization will equalize variation in smoking history and other variables between the two study arms, thus ensuring high quality data. JUUL e-cigarette with tobacco flavoured 5% nicotine strength pods have been chosen because of their impressive nicotine delivery profile (Yingst et al., 2019; Hajek P et al., 2020; Maloney et al., 2020), the reported high quit rates in smokers with schizophrenia spectrum disorders (Caponnetto et al., 2018b) and - last but not least – because they are very easy and safe to use (the pod system design does not require manual re-filling – an important safety aspect when considering that the target population in this study is people with schizophrenia). As it is unethical not to provide some nicotine substitution for such highly dependent population, the reference group will be randomized to a lower strength of nicotine (1.5%). Of note, 5% and 1.5% nicotine pods are identical in appearance, and therefore suitable for double-blinding. When comparing quit rates between the two nicotine strengths (5% vs. 1.5%) it will be also possible to gather useful findings that may inform future regulatory decisions in relation to restrictions about nicotine levels in vaping products (e.g see Tobacco Products Directive).

The protocol incorporates a number of innovative approaches that contribute to the specific uniqueness and quality of the study. A prospective monitoring of cigarette consumption and e-cigarette use will be carried out throughout the study; participants will self-report cigarette consumption and e-cigarette usage at each study visit in their study diary. Moreover, subjects will be asked to return all empty, part-used, and unused consumables (i.e. e-cigarette pods). Any non-compliance will be recorded in the study diary after counting all empty, part-used and unused pods returned at each visit. Self-reported cigarette and/or e-cigarette use will be also constantly monitored in their Tracker App, which will also allow recording and verification of any non-compliance with the protocol as well as participants’ self-rated mental health status. Participants will also be informed that biochemical verification of compliance as well as assessments of adherence will be conducted at each clinic visit. Of note, non-compliance to study products is in itself an interesting outcome and we will be able to assess the impact of different level of non-compliance on smoking cessation rates. Trial attendance and retention of the participants is expected to be high given that participants have to return to the clinic for their regular re-supply of e-cigarette pods. Another unique aspect of the study design is the inclusion of the 6-moths extension period to investigate changes in vaping/smoking behavior after withdrawal of study products (i.e. continuous abstainance, smoking relapse) under naturalistic condition, as well as to review tolerability and THR potential in the longer term.

We foresee the following potential limitations and challenges; 1) study findings will be product specific and likely to be limited in generalizability to other vaping products; 2) evolution of the COVID-19 pandemic will be still unpredictable and likely to slow down recruitment and completion of the study if impacting participating sites.

Engaging this vulnerable tobacco dependant population with schizophrenia spectrum disorders has immense consequences for the reductions of overall burden of smoking globally. Results from the study may contribute to a better understanding of e-cigarettes as a less harmful alternative to tobacco smoking with the possibility of health gains in smokers with schizophrenia spectrum disorders.

## Data Availability

NA

## LIST OF ABBREVIATIONS

AE: Adverse Event
ANOVA: One-way analysis of variance
APR: Annual progress report
AR: Adverse Reaction
BP: Blood pressure
CI: Chief Investigator
CO: Carbon Monoxide in exhaled breath
CRF: Case Report Form
CRO: Contract Research Organisation
CST: Chester step test
DMC: Data Monitoring Committee
EQ VAS: Quality of Life scale
FTCD: Fagerstrom test for cigarette dependence
GCP: Good Clinical Practice
HR: Heart rate
IB: Investigator Brochure
mCEQ: Modified Cigarette Evaluation Questionnaire
PI: Principal Investigator
REC: Research Ethics Committee
SAE: Serious Adverse Event
PANSS: Positive and negative symptoms of Schizophrenia
SAR: Serious Adverse Reaction
SmPC: Summary of product characteristics
SOP: Standard Operating Procedure
SRMH: Self-rated mental health
SUSAR: Suspected Unexpected Serious Adverse Reaction
TC: Telephone Contact
TSC: Trial Steering Committee

## TRIAL FLOW CHART

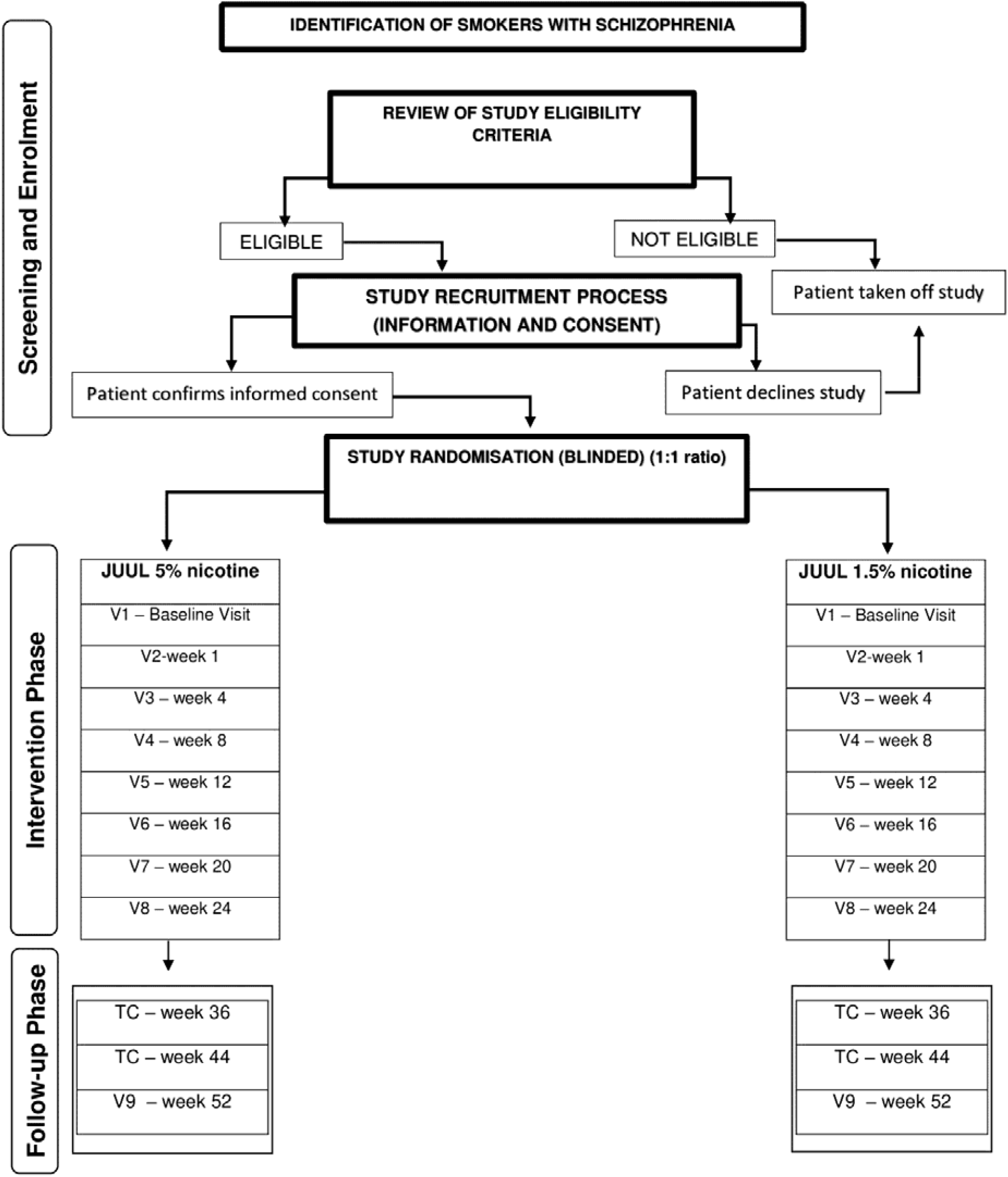

